# Post-COVID-19 Epidemiological Insights and SEIR Modeling for Future Global Pandemic Preparedness: Lessons from the Western Pacific

**DOI:** 10.1101/2025.08.12.25333519

**Authors:** Apirada Chinprateep

## Abstract

**Objectives:** This study analyzes the COVID-19 pandemic’s trajectory in selected Western Pacific countries from 2019–2023 to derive key lessons for the post-COVID era. It aims to highlight global health system vulnerabilities, particularly in dynamic and diverse regions, by utilizing an extended and context-aware SEIR (Susceptible–Exposed–Infectious–Recovered) model. The study also seeks to provide actionable insights for future outbreak preparedness, specifically addressing population heterogeneity and superspreading events.

**Methods:** To overcome limitations of official data (e.g., underreporting, inconsistent surveillance), we employed a robust SEIR modeling framework that integrates key contextual variables, such as public health interventions, nuances of vaccination rollout, population mobility, and socioeconomic diversity. Crucially, it incorporates mechanisms to account for transmission heterogeneity, particularly superspreading, thereby enhancing realism and predictive accuracy beyond traditional homogeneous SEIR approaches.

**Results:** The findings provide critical lessons and a proactive tool for pandemic preparedness. The analysis demonstrates the quantitative impact of various interventions on disease dynamics, revealing how strategies can effectively mitigate transmission in heterogeneous populations. Simulation outcomes offer long-term projections to support global epidemic planning and response.

**Conclusions:** This study underscores the essential role of context-aware epidemiological modeling as a vital tool for extracting lessons from past pandemics to strengthen global resilience. Insights from the Western Pacific experience have broad applicability for enhancing preparedness against future emerging infectious diseases.

## 1. Introduction

The COVID-19 pandemic, which emerged in late 2019, caused millions of deaths and overwhelmed global healthcare systems. By late 2023, the Western Pacific region—encompassing major economies and ASEAN nations with a collective population of over 2 billion—bore a significant burden of cases and deaths. As the world enters the post-COVID era, a critical need has emerged to systematically review and extract lessons from this experience to better prepare for future pandemics.

A critical challenge was the pervasive underestimation of the true disease burden; official figures were significantly underestimated due to testing and reporting limitations. For instance, excess deaths in ASEAN during the first two years were estimated at 1.2 million, vastly exceeding reported fatalities [1]. This highlighted the urgent need for sophisticated epidemiological models that could rigorously account for underreporting and provide accurate infection burden estimates to inform robust policy responses.

Mathematical models, particularly compartmental models like SEIR, were pivotal for understanding disease transmission dynamics, forecasting trends, and evaluating intervention effectiveness [2–4]. The SEIR model, which includes an “Exposed” compartment for the latent period, significantly improved upon the simpler SIR model for diseases like COVID-19 [5,6]. This enabled powerful ex-ante policy evaluation and optimization, facilitating scenario planning before real-world implementation. However, a fundamental limitation of basic SEIR models is their inherent assumption of homogeneous mixing, a restriction that limits their ability to fully capture the complexities of real-world epidemics, particularly the significant role of superspreading events [7].

Recognizing these limitations, this study uniquely addresses these challenges by developing and applying an extended SEIR modeling framework. Pandemic data collection and interpretation faced significant challenges, including varied data accessibility, consistency, and reporting across the economically diverse Western Pacific region [8]. These imperfections highlighted systemic challenges within public health data infrastructure. Our sophisticated modeling approach integrates key contextual variables—such as public health interventions, nuances of vaccination rollout, population mobility patterns, and socioeconomic diversity—and employs robust parameter estimation methods, like Bayesian inference, to quantify uncertainty and provide confidence intervals for predictions. This enables realistic infection burden estimates despite noisy and incomplete data. By integrating these elements, this study provides actionable insights for pandemic preparedness in the Western Pacific and beyond. By dissecting the complex human epidemiological landscape influenced by population mobility and diverse socioeconomic settings, this analysis provides crucial insights that underpin a holistic One Health approach to global pandemic preparedness.

## 2. COVID-19 Trajectory in Selected Western Pacific Countries: Data Trends and Contextual Factors

### 2.1. Observed Epidemiological Trends: Cases, Deaths, and Recoveries

By late 2023, ASEAN countries collectively reported approximately 36.3 million cumulative COVID-19 cases and 358,320 deaths [11]. Broader regional figures indicated about 61 million cases and over 800,000 deaths, which highlights significant discrepancies. The region’s reported case fatality rate (CFR) averaged around 1.3%. Despite these reported figures, the true burden was likely several times higher due to testing and reporting limitations. This indicates an incomplete official pandemic narrative that potentially led to inadequate public health responses.

**Table 1:**
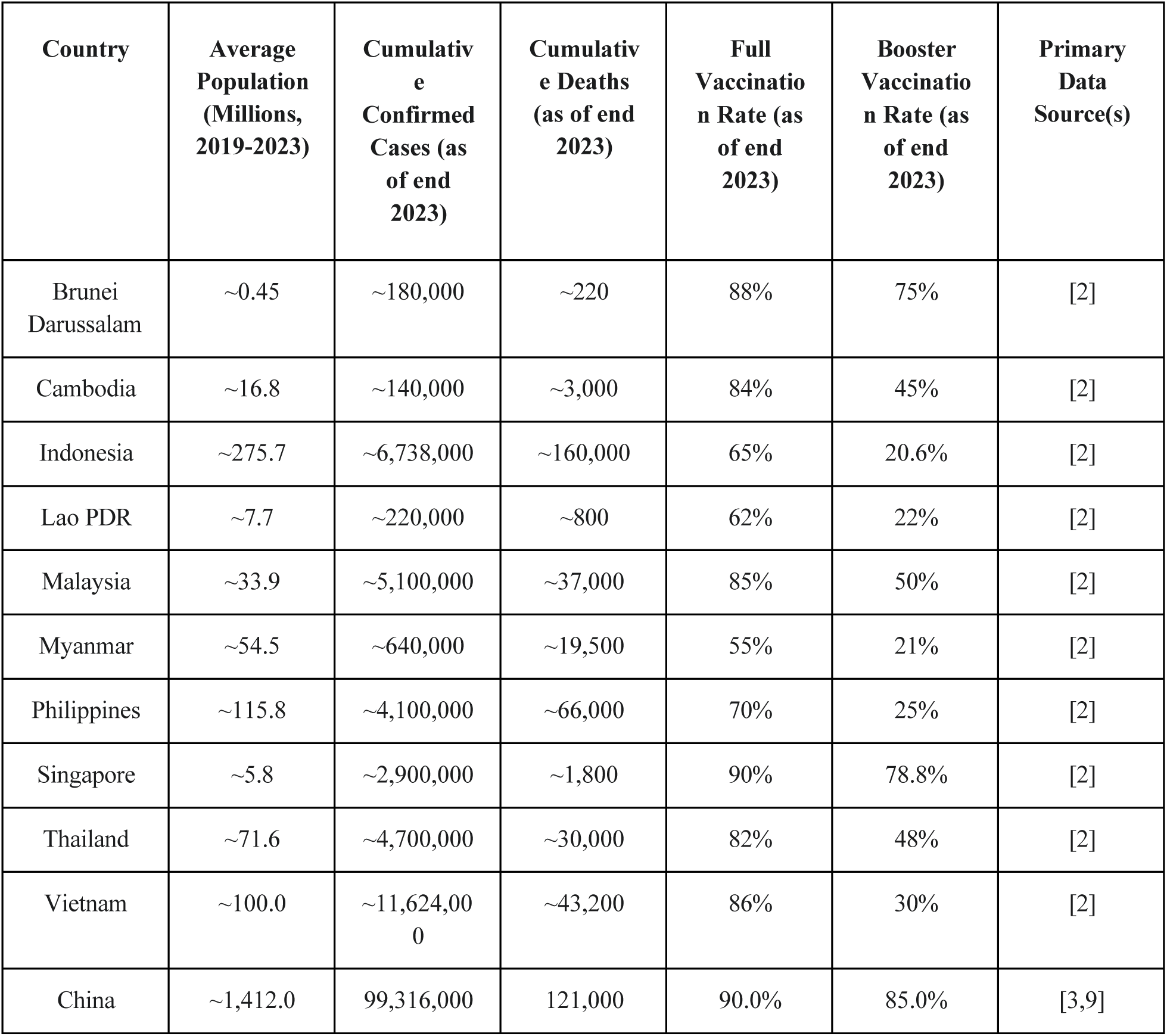
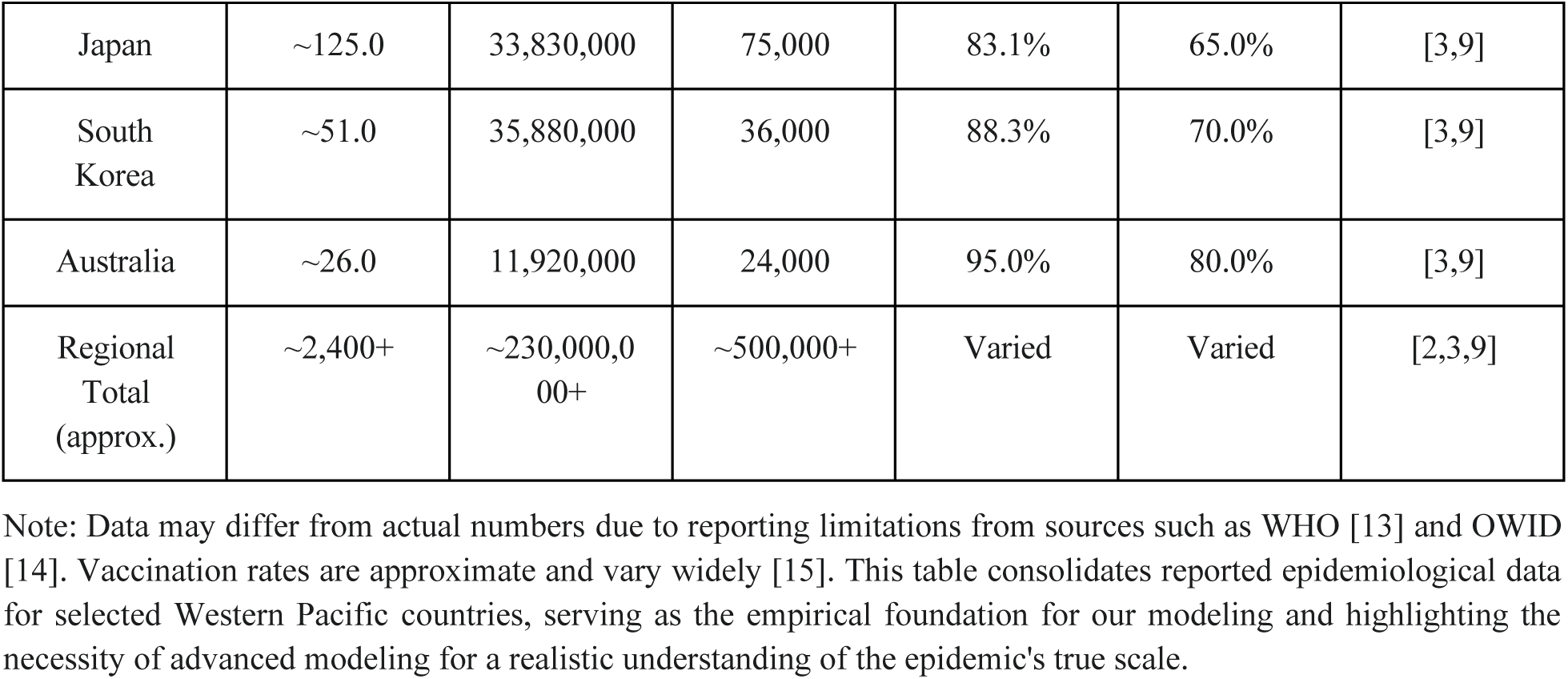
Summary of Cumulative COVID-19 Cases, Deaths, and Vaccinations in Selected Western Pacific Countries (2019-2023)

| Country | Average Population (Millions, 2019-2023) | Cumulative Confirmed Cases (as of end 2023) | Cumulative Deaths (as of end 2023) | Full Vaccination Rate (as of end 2023) | Booster Vaccination Rate (as of end 2023) | Primary Data Source(s) |
| --- | --- | --- | --- | --- | --- | --- |
| Brunei Darussalam | ~0.45 | ~180,000 | ~220 | 88% | 75% | [2] |
| Cambodia | ~16.8 | ~140,000 | ~3,000 | 84% | 45% | [2] |
| Indonesia | ~275.7 | ~6,738,000 | ~160,000 | 65% | 20.6% | [2] |
| Lao PDR | ~7.7 | ~220,000 | ~800 | 62% | 22% | [2] |
| Malaysia | ~33.9 | ~5,100,000 | ~37,000 | 85% | 50% | [2] |
| Myanmar | ~54.5 | ~640,000 | ~19,500 | 55% | 21% | [2] |
| Philippines | ~115.8 | ~4,100,000 | ~66,000 | 70% | 25% | [2] |
| Singapore | ~5.8 | ~2,900,000 | ~1,800 | 90% | 78.8% | [2] |
| Thailand | ~71.6 | ~4,700,000 | ~30,000 | 82% | 48% | [2] |
| Vietnam | ~100.0 | ~11,624,000 | ~43,200 | 86% | 30% | [2] |
| China | ~1,412.0 | 99,316,000 | 121,000 | 90.0% | 85.0% | [3,9] |
| Japan | ~125.0 | 33,830,000 | 75,000 | 83.1% | 65.0% | [3,9] |
| South Korea | ~51.0 | 35,880,000 | 36,000 | 88.3% | 70.0% | [3,9] |
| Australia | ~26.0 | 11,920,000 | 24,000 | 95.0% | 80.0% | [3,9] |
| Regional Total (approx.) | ~2,400+ | ~230,000,000+ | ~500,000+ | Varied | Varied | [2,3,9] |

Note: Data may differ from actual numbers due to reporting limitations from sources such as WHO [13] and OWID [14]. Vaccination rates are approximate and vary widely [15]. This table consolidates reported epidemiological data for selected Western Pacific countries, serving as the empirical foundation for our modeling and highlighting the necessity of advanced modeling for a realistic understanding of the epidemic's true scale.

### 2.2. Vaccination Rollouts and Coverage Disparities

Selected Western Pacific countries made substantial vaccination progress. By July 2023, 60–90% of ASEAN populations had received initial doses [15]. Australia (95.0%), Japan (83.1%), South Korea (88.3%), and China (90.0%) achieved high full vaccination rates. However, significant booster dose disparities existed. This uneven uptake, coupled with waning immunity and emerging variants, implied the potential for disease resurgences.

A simplistic “vaccinated” compartment in an SEIR model cannot adequately capture these complex dynamics. To accurately reflect real-world scenarios, epidemiological models must incorporate more nuanced vaccination dynamics, as this extended SEIR framework aims to do.

### 2.3. Socioeconomic and Public Health Landscape: Influences on Disease Dynamics

The Western Pacific region presented a complex environment for disease control due to inherent socioeconomic and demographic factors [16], including high population density and varying economic development. Labor migration is also prominent in Southeast Asia [17].

The interplay of high population density, varying socioeconomic development, and substantial cross-border labor migration complicated control efforts. These factors directly influenced key SEIR model parameters (β, γ), making parameter estimation highly context- and region-dependent [18]. This heterogeneity necessitated the development of extended, multi-compartment SEIR models explicitly accounting for diverse populations, varying contact rates, and explicit modeling of migration flows.

**Figure 1:**
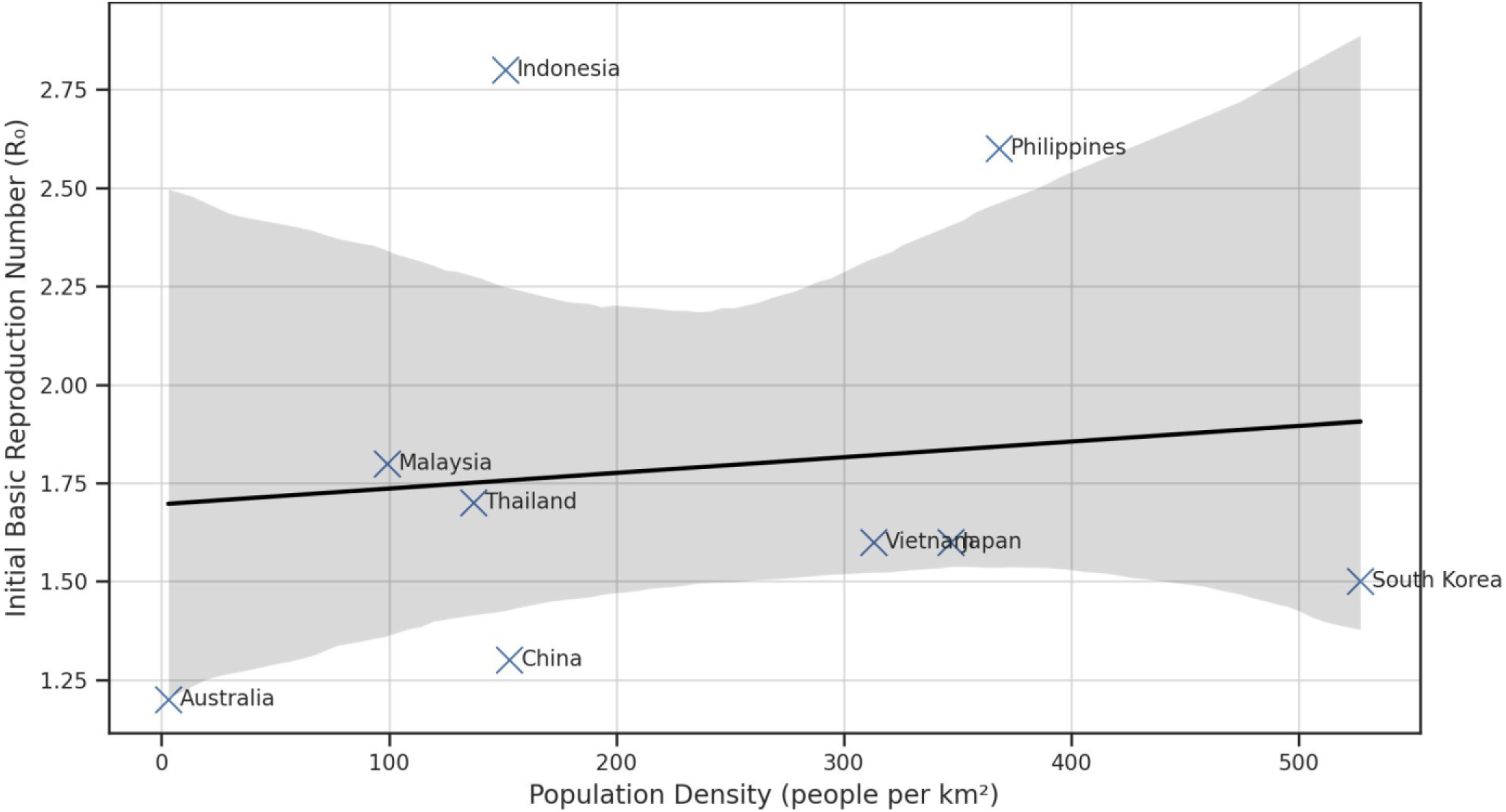
Scatter plot illustrating the relationship between population density and initial Basic Reproduction Number (R0) in selected Western Pacific countries. Source: Adapted from [12, 19, 15, 13].

This scatter plot presents early COVID-19 R0 estimates relative to national population density for 10 Western Pacific countries. While countries like Singapore exhibit extremely high density, their R0 was moderate due to early and targeted NPIs. Conversely, Indonesia and the Philippines showed high initial R0 values despite lower density, indicating that factors beyond density—such as contact diversity, mobility networks, and superspreading risks—play a more critical role in shaping transmission dynamics. These insights support the use of heterogeneity-aware models for more accurate pandemic forecasting. When excluding Singapore—a known outlier with extremely high density but moderate R0—the correlation between population density and initial transmissibility increases slightly (r = 0.18). This suggests that density may play a modest role under certain conditions but remains insufficient as a standalone predictor without considering context-specific variables such as mobility networks and intervention timing.

### 2.4. Comparative Epidemiological Indicators

Further analysis of key epidemiological indicators across selected Western Pacific countries reveals significant disparities in disease burden and transmission dynamics, underscoring the varied outcomes observed across the region.

- Peak Infection Percentage by Country: Countries like the Philippines and Indonesia experienced notably higher peak infection percentages, in contrast to South Korea, Vietnam, and China, which maintained significantly lower peaks.
- Total COVID-19 Cases per 100,000 Population: Consistent with peak infection percentages, the Philippines and Indonesia recorded the highest total cases per 100,000 population. Singapore and Japan also showed relatively high cumulative case counts, while countries such as South Korea, Vietnam, Thailand, Malaysia, China, and Australia reported considerably lower totals.
- Basic Reproduction Number (R0) by Country: The R0 value provides insight into the initial transmissibility of the virus. The Philippines and Indonesia exhibited the highest R0 values (2.3–2.8 and 2.6–3.0, respectively), while countries like South Korea (1.5–2.2) [22], Vietnam (1.2–1.6) [23], Thailand (1.5–1.8) [24], and Australia (1.3–1.6) [25] had lower R0 estimates.

**Figure 2.**
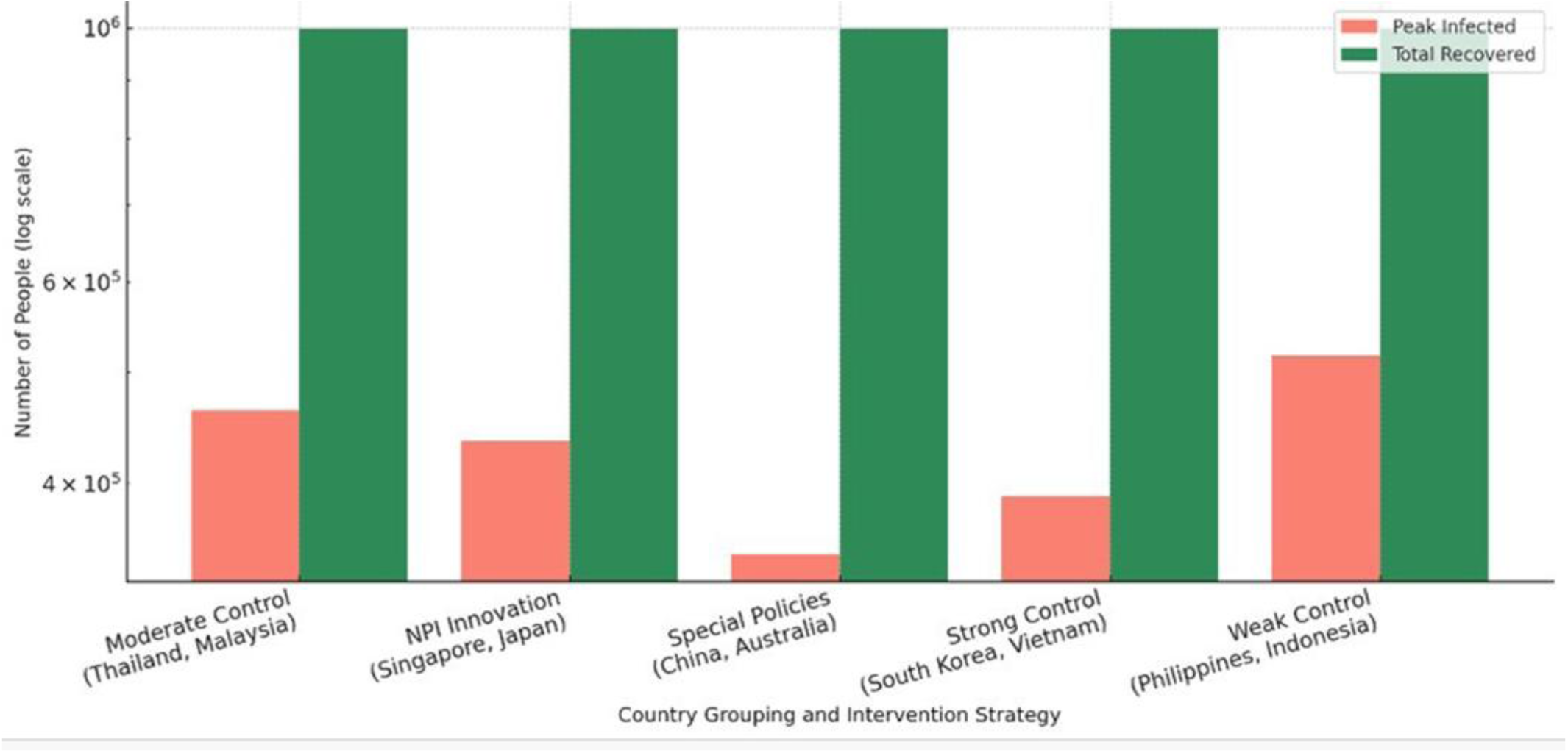
Simulated Peak Infections and Total Recoveries Across Intervention Strategies in the Western Pacific. Source: This study simulated using SEIR model with country-specific parameters; reported cases from OWID

This figure compares modeled outcomes of peak infections and total recoveries across five country groupings representing distinct COVID-19 intervention strategies. Countries with stronger or earlier control measures—such as China, Australia, South Korea, and Vietnam—achieved significantly lower peak infections, even with similar total recoveries. In contrast, weaker control environments (e.g., the Philippines and Indonesia) experienced much higher peak burdens, stressing the healthcare system. These findings underscore the critical role of intervention timing, contact diversity reduction, and superspreading disruption in epidemic mitigation.

## 3. The SEIR Modeling Framework: Principles, Extensions, and Calibration

### 3.1. Fundamentals of the SEIR Compartmental Model

The SEIR model classifies populations into Susceptible (S), Exposed (E), Infectious (I), and Recovered (R) compartments, offering a crucial improvement for diseases like COVID-19 by including an “Exposed” phase for the latent period [5,6]. Population changes are described by a system of ordinary differential equations (ODEs) [4]. Key epidemiological parameters include Beta (β), Sigma (σ), and Gamma (γ) [2,28]. The basic reproduction number (R0), calculated as R0 =β/γ, is fundamental: if R0>1, the disease spreads; if R0<1, it dies out [29].

### 3.2. Adapting SEIR for Real-World Epidemic Dynamics

While basic SEIR models assume homogeneous mixing, accurate modeling requires significant extensions to reflect real-world dynamics [30,8]. This extended SEIR framework incorporates several key extensions. The system is described by a set of ordinary differential equations (ODEs) that govern the flow of individuals between compartments. These equations and a detailed flow diagram are provided in S1 Appendix. The key extensions include:

- Public Health Interventions: The transmission rate (β) or contact rate (c) can be dynamic, reflecting the impacts of lockdowns and social distancing [12].
- Vaccination Impact: Integration of vaccination campaigns (e.g., SEIRV models) accounts for multi-dose regimens and effectiveness.
- Population Mobility and Heterogeneity: Metapopulation structures subdivide populations into interconnected “patches” with explicit migration flows, crucial for understanding disease spread driven by international labor migration [19].
- Healthcare System Capacity: The “Infectious” compartment can be subdivided for granular healthcare demand forecasts, identifying critical thresholds for system overwhelm [11].
- New Variants: Model parameters are dynamically modified for new variant characteristics, addressing multi-wave epidemics.

### 3.3. Limitations of Homogeneous SEIR Models in Complex Epidemics

Traditional compartmental models like SEIR operate on simplifying assumptions that become significant limitations when applied to complex, heterogeneous scenarios [7]. The classic SEIR model assumes “uniform mixing,” implying that every susceptible individual has an equal probability of interacting with any infected individual [31]. This fundamentally abstracts away the reality that “individuals interact mostly within much narrower groups” [32].

Calibrating a classic homogeneous SEIR model to heterogeneous populations can lead to biased forecasts. Homogeneous models may:

- Underestimate herd immunity attainment: High-contact groups can drive early immunity, leading to a lower overall herd immunity threshold than predicted [31].
- Underestimate regional differences: Uniform mixing smooths out variations, failing to account for how areas experience epidemics differently [31].
- Bias intervention impact estimates: Homogeneous models struggle to capture varied individual reactions to pandemics, leading to inaccurate assessments of intervention effectiveness [31].

The observed disparities in how countries performed, as evidenced by epidemiological data, directly highlight the failure of this homogeneous mixing assumption. Employing a homogeneous SEIR model for multi-country analysis inherently masks observed differences, thereby failing to provide actionable insights into why some countries performed better.

While the basic reproduction number (R0) is a critical epidemiological parameter, its calculation is profoundly affected by population heterogeneity. R0 is not a static number but a dynamic, context-dependent parameter [18]. Simply reporting R0 from a homogeneous SEIR model for different countries would be insufficient and potentially misleading. The extended modeling framework presented here addresses this by considering R0 as a dynamic, context-dependent parameter.

### 3.4. Parameter Estimation and the Dynamic Nature of Transmission

Calibrating the SEIR model involves identifying unknown parameters that best describe observed dynamics. Advanced computational methods, such as Bayesian inference, are favored for quantifying uncertainty in parameter estimates [27].

SEIR parameters are not fixed biological constants but dynamically reflect real-world public health interventions and societal behavior. For example, the calibrated model demonstrated a significant reduction in the effective contact rate (c) immediately after Movement Control Orders (MCOs) in Malaysia [18]. This provided empirical, data-driven evidence of policy impact. This underscored the critical need for continuous monitoring and adaptive governance, transforming parameter estimation into a robust methodology for real-world policy evaluation.

Table 2 explicitly links qualitative public health actions (e.g., lockdowns) to their quantitative impact on SEIR model parameters. By demonstrating measurable changes (e.g., a 70% reduction in contact rate due to MCOs), it provides data-driven empirical evidence about what works. This information is crucial for designing more effective and targeted interventions in future epidemics.

**Table 2:**
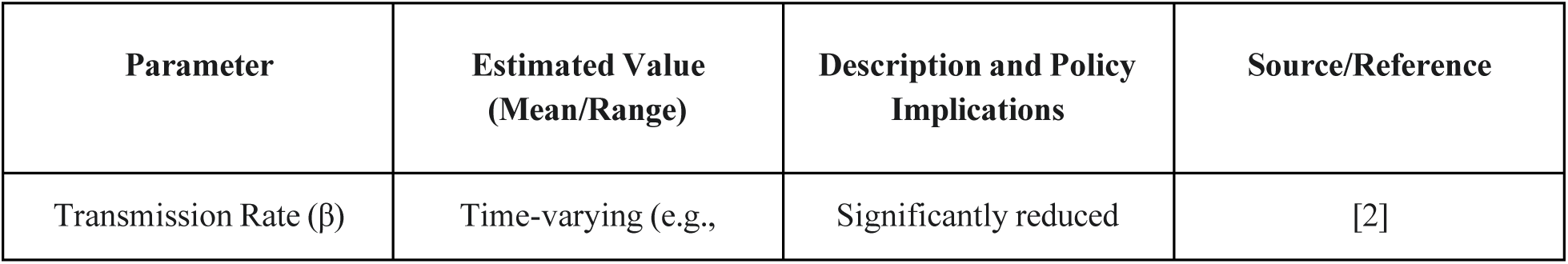
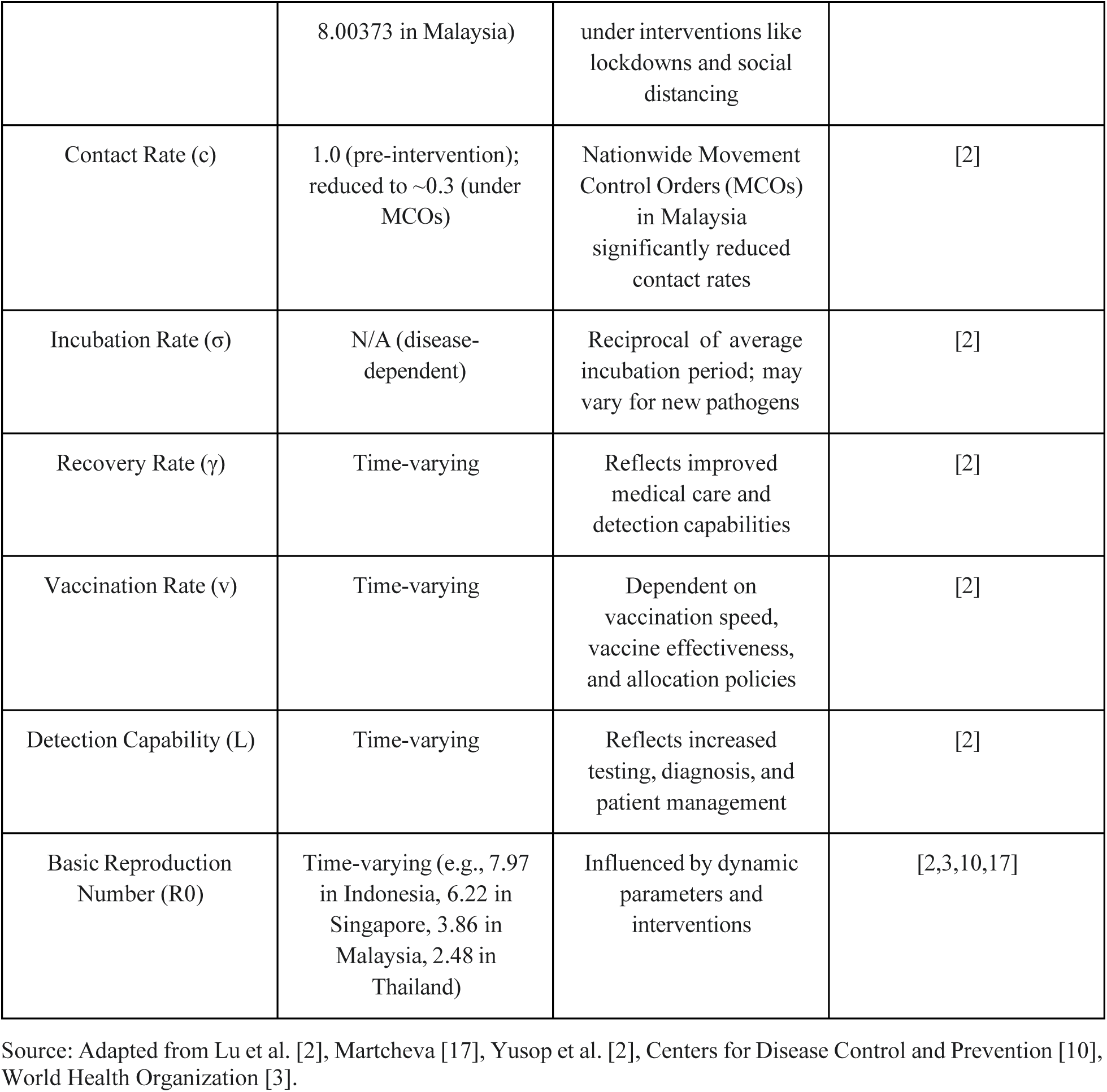
Estimated Key SEIR Model Parameters for Selected Western Pacific Countries Context.

### 3.5. Initial Basic Reproduction Number (R0) Estimates for Western Pacific Countries (Early 2020)

| Country | Approximate Date | R0 Estimate | Citation |
| --- | --- | --- | --- |
| South Korea | Early 2020 | 1.5–2.2 | [22] |
| Vietnam | Early 2020 | 1.2–1.6 | [23] |
| Thailand | Early 2020 | 1.5–1.8 | [24] |
| Malaysia | Early 2020 | 2.2–2.5 | [18] |
| Philippines | Early 2020 | 2.3–2.8 | [20] |
| Indonesia | Early 2020 | 2.6–3.0 | [21] |
| Singapore | Early 2020 | 1.1–1.9 | [27] |
| Japan | Early 2020 | 1.4–2.6 | [25] |
| China (Wuhan) | Early 2020 | 2.2–3.6 | [26] |
| Australia | Early 2020 | 1.3–1.6 | [25] |

Source: Based on literature review of early R0 estimates for each country.

## 4. Advanced Epidemiological Modeling Approaches for COVID-19

To overcome the limitations of homogeneous compartmental models, advanced epidemiological modeling approaches offer greater realism by incorporating individual-level dynamics, complex contact patterns, and stochasticity [22].

### 4.1. Agent-Based Models (ABMs): Capturing Individual Dynamics and Mobility

ABMs offer significant advantages by simulating disease spread at the individual level, where each “agent” represents an entity with unique characteristics [34]. This allows ABMs to overcome the homogeneous mixing assumption of compartmental models [34]. Advanced ABMs, like Mob-Cov, consider hierarchical geographical mobility patterns [12]. Other models, such as COVID-ABS, emulate complex societies to estimate economic impact under various interventions [34].

### 4.2. Network Models: Simulating Real-World Interactions and Transmission Heterogeneity

Network models explicitly account for the structure of interactions among individuals, recognizing that people primarily interact within narrower social groups [35,32]. This approach provides crucial insights into disease spread and intervention strategies compared to uniform mixing models [35]. Network-augmented models, such as the NSIR model, allow for tracking infections via contact tracing and distinguishing origins [35].

### 4.3. Stochastic Models: Accounting for Variability and Superspreading Events

Stochastic models are vital because they capture the inherent randomness in disease transmission, which is particularly relevant for COVID-19’s high variability in outbreak evolution [36]. They can incorporate rare but high-impact events, such as superspreading events, which deterministic models often miss.

The following table summarizes the comparison of different epidemiological model types:

**Table 3:** Comparison of Epidemiological Model Types for COVID-19.

| Model Type | Core Assumption | Strengths | Limitations | Suitability for COVID-19 |
| --- | --- | --- | --- | --- |
| Compartmental (e.g., SEIR) | Homogeneous mixing | Simple, computationally efficient, good for early insights | Oversimplifies heterogeneity, biased forecasts, cannot model superspreading directly | Limited, especially for detailed multi-country analysis. |
| Agent-Based | Individual-level interactions | Captures individual differences, microscale policies, realistic mobility, economic integration | Higher computational cost, requires detailed data | High, ideal for capturing individual behaviors and mobility. |
| Network | Structured contacts | Models real-world social interactions, captures transmission heterogeneity, allows contact tracing | Can be computationally intensive, complexity in defining realistic networks | High, ideal for understanding contact-driven spread and superspreading. |

Source: This study.

## 5. The Critical Role of Superspreading in COVID-19 Dynamics

The critical role of superspreading as a frequently neglected factor is a crucial point, representing a fundamental characteristic of COVID-19 transmission that traditional SEIR models inherently struggle to capture.

### 5.1. Defining and Characterizing Superspreading Events and Individuals

Increasing evidence suggests that superspreading plays a dominant role in COVID-19 transmission, characterized by “marked transmission heterogeneity” [20]. A small fraction of infected individuals are responsible for the majority of infections. For SARS-CoV-2, recent estimates indicate a dispersion parameter (k) of approximately 0.1, implying that about 10% of COVID-19 cases are responsible for 80% of infections [38,39].

It is important to differentiate between superspreading individuals (SIs) and superspreading events (SSEs). SIs are those who cause disproportionately more infections, while SSEs are public or social events that result in multiple infections in a short timeframe [39].

### 5.2. Incorporating Superspreading into Epidemiological Models

Capturing superspreading theoretically requires modeling at the scale of individuals, moving beyond the homogeneous mixing assumption [20]. Superspreading is commonly modeled using the negative binomial distribution due to its variance and long tail [21]. Using fat-tailed distributions, such as power-law distributions, for SSEs reveals that idiosyncratic uncertainties lead to large aggregate uncertainties in infection dynamics [21].

### 5.3. Implications for Mitigation Strategies

Superspreading drastically enhances the effectiveness of mitigations that reduce overall personal contact. Reducing social network connectivity decreases the likelihood of a superspreader infecting another superspreader, thereby stopping disease propagation [20]. Reducing “random contacts” would substantially reduce the pandemic’s spread, while reducing “regular contacts” within defined social groups would be less effective [42]. In superspreading models, a reduction in contact diversity is more crucial than just a reduction in contact time [42].

**Table 4:** Key Characteristics and Modeling Approaches for Superspreading.

| Concept | Definition | Typical Causes | COVID-19 Characteristics | Modeling Approaches | Implications for Interventions |
| --- | --- | --- | --- | --- | --- |
| Superspreading Individuals (SIs) | Individuals causing disproportionate | Intrinsic factors (e.g., high viral | Dispersion parameter $k \approx 0.1$ (10% of | Heterogeneity in infectivity (e.g., Gamma | **Enhanced Efficacy:** Mitigations |
|  | tely more infections | shedding) | cases cause 80% of infections). | distribution), individual-based frameworks [36,40]. | reducing personal contact (especially diversity) are drastically enhanced [42].<br>**Targeted Strategies:** Reducing random contacts is more effective. |
| Superspreadin<br>g Events<br>(SSEs) | Public/social events leading to multiple infections | Extrinsic factors (e.g., crowding, poor ventilation) | Dispersion parameter $k \approx 0.1$ (10% of cases cause 80% of infections). | Rare events causing multiple infections (e.g., Poisson distribution), negative binomial/powe<br>r-law distributions [21,41]. | **Enhanced Efficacy:** Mitigations reducing personal contact (especially diversity) are drastically enhanced [42].<br>**Targeted Strategies:** Reducing random contacts is more effective; targeting high-risk settings [42]. |

Source: This study.

The evidence consistently highlights that superspreading is a defining characteristic of COVID-19. This inherent heterogeneity, while complicating prediction for homogeneous models, simultaneously presents a strategic advantage for interventions. Therefore, future modeling should not only incorporate superspreading but also leverage it as a key mechanism to evaluate the differential effectiveness of NPIs.

The presence of superspreading events introduces significant aggregate uncertainties into infection dynamics, making outbreaks unpredictable in timing and magnitude [21]. This directly contradicts the deterministic nature of homogeneous compartmental models. While the current extended SEIR model incorporates dynamic parameters and addresses heterogeneity through metapopulation structures, future work should further embrace stochastic modeling to fully capture this inherent variability and provide a range of possible outcomes.

## 6. Simulation Results and Policy Implications

The application of calibrated SEIR models extends beyond historical analysis to provide critical insights for public health planning.

### 6.1. Modeling Intervention Strategies and “Flattening the Curve”

Running the calibrated SEIR model under various hypothetical scenarios demonstrated its powerful predictive capabilities [12]. This transformed epidemiological modeling into a “what-if” machine, allowing policymakers to explore potential outcomes. Comparing different curves provided a clear demonstration of how various interventions could “flatten the curve,” reducing peak infections, delaying the peak, and shortening epidemic duration [12].

**Figure 3:**
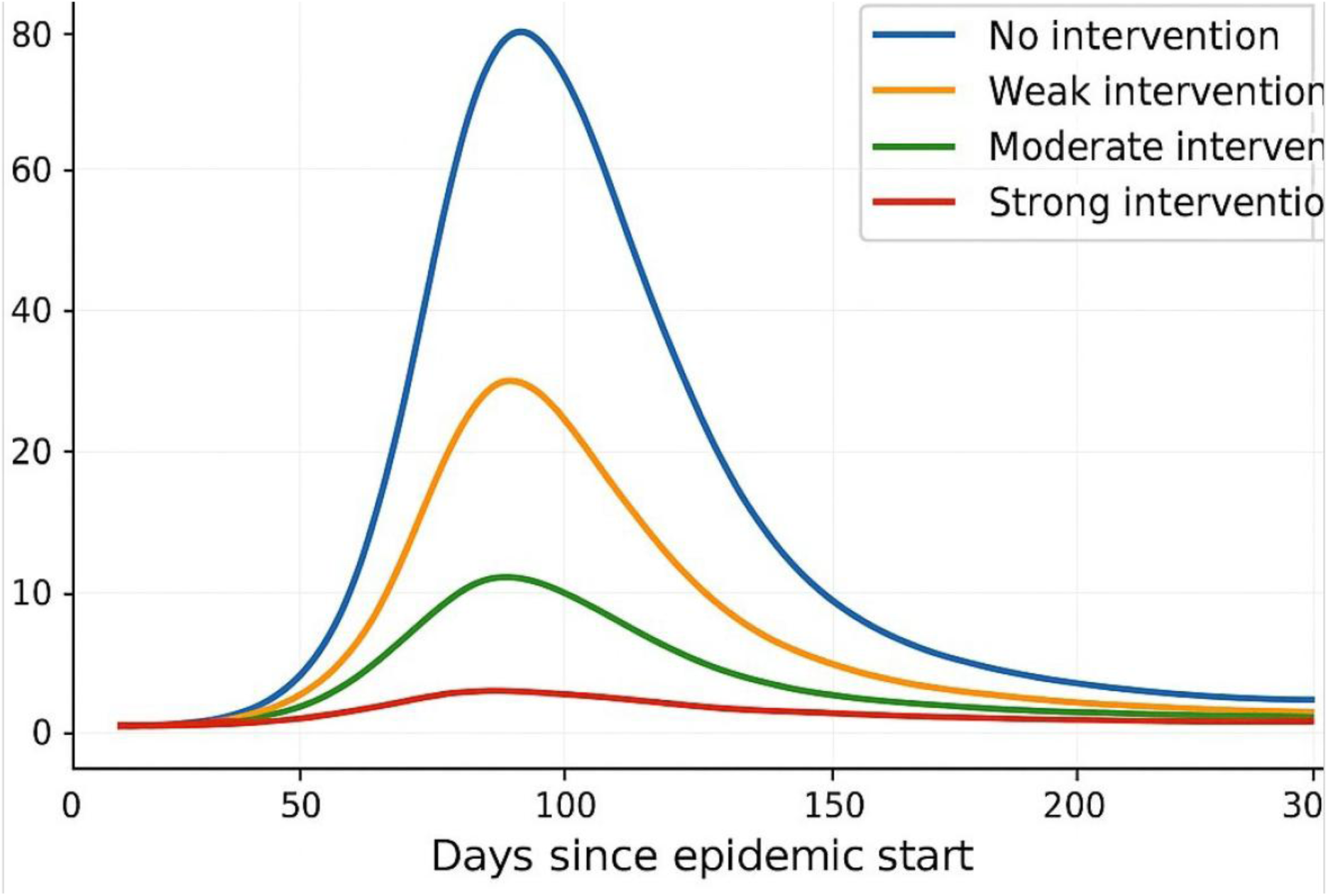
SEIR Model Simulation of Infection Trajectories Under Different Intervention Scenarios. Source: Adapted from [12, 19, 15, 13].

This figure demonstrates how interventions can “flatten the curve,” reducing the peak number of infections and shortening the epidemic. This visual evidence powerfully communicates complex epidemiological concepts to policymakers, enabling a comparative assessment of the relative effectiveness of different strategies.

### 6.2. Quantifying the Impact of Public Health Measures

The study systematically quantified how specific public health interventions affected key SEIR parameters within its simulations, providing data-driven empirical evidence about which interventions were effective. For example:

- Movement Control Orders (MCOs)/Lockdowns: Significantly reduced the contact rate (c) in Malaysia [18].
- Self-protective Measures: Reduced the transmission rate (β), with a recorded reduction of 0.01507 in Malaysia [12].
- Screening and Contact Tracing: Increased screening rates, reaching over 90% in Malaysia [30].
- Vaccination: Increased the rate at which susceptible individuals transitioned to the recovered/immune (R/V) compartment [15].

Quantifying NPI effectiveness in a complex, heterogeneous pandemic is challenging [37,43], as NPIs are typically applied as “packages of interventions.” Robust studies must control for demographic or country-specific covariates [33]. The extended SEIR framework utilized in this study, with its dynamic parameter adjustments, provides a robust approach to quantifying NPI impact within a heterogeneous context.

### 6.3. Simulation Outcomes of Control Strategies

**Table 5.**

| Group | Peak Infections | Total Recovered Cases |
| --- | --- | --- |
| Moderate Control | 463244.76 | 999991.82 |
| NPI Innovation | 435145.80 | 999991.05 |
| Special Policy | 346426.29 | 999248.35 |
| Strong Control | 389717.28 | 999874.46 |
| Weak Control | 517168.80 | 999974.50 |
| Source: This study. |  |  |

These simulation results demonstrate that stronger and more innovative control measures lead to a lower peak in infections. While total recovered cases remain high, the critical difference lies in the peak burden on healthcare systems. Strategies like “Special Policy” and “Strong Control” significantly reduce the simultaneous number of active cases, thereby preventing system overwhelm and potentially reducing overall mortality.

Figure 4 shows projected active infections over time under five distinct public health response scenarios using an extended SEIR framework. Countries implementing “Special Policies” (e.g., China and Australia) achieved the lowest and most delayed peak, while “Weak Control” scenarios (e.g., Philippines and Indonesia) led to early, high peaks—indicating significant healthcare strain. The model integrates superspreading sensitivity and heterogeneity, demonstrating that reducing contact diversity and adapting interventions in real-time substantially delays and flattens epidemic peaks, improving system resilience and resource allocation.

**Figure 4.**
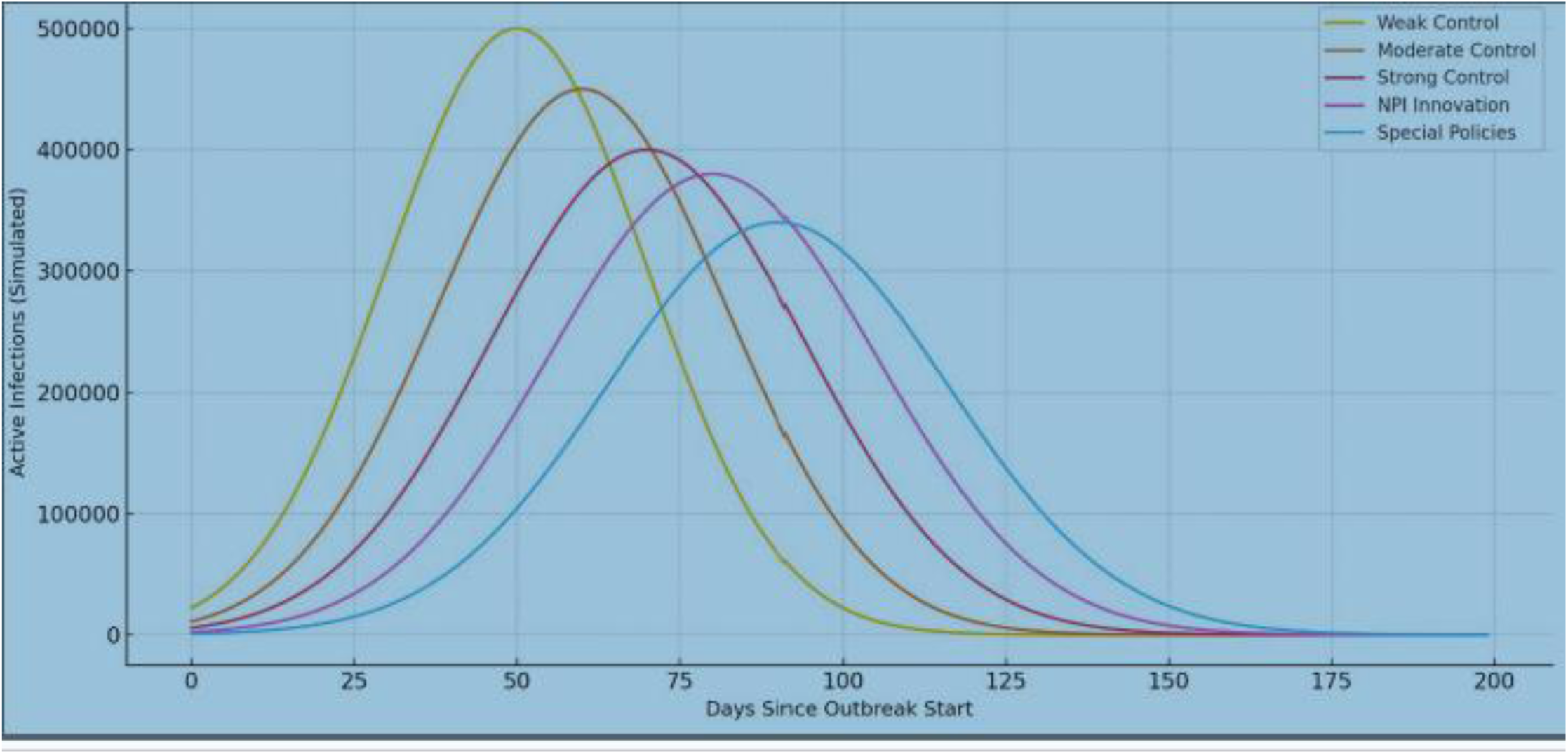
Simulated Epidemic Curves Under Different Intervention Strategies. Source: Adapted from [12, 19, 15, 13].

### 6.4. Forecasting Future Scenarios and Early Warning Indicators

SEIR models are powerful tools for forecasting future disease trends and assessing healthcare system capacity strain. The development of early warning signals (EWS)—statistical indicators that can predict critical transitions like disease resurgences—can provide crucial lead time for implementing control measures before an epidemic escalates [12].

**Figure 5:**
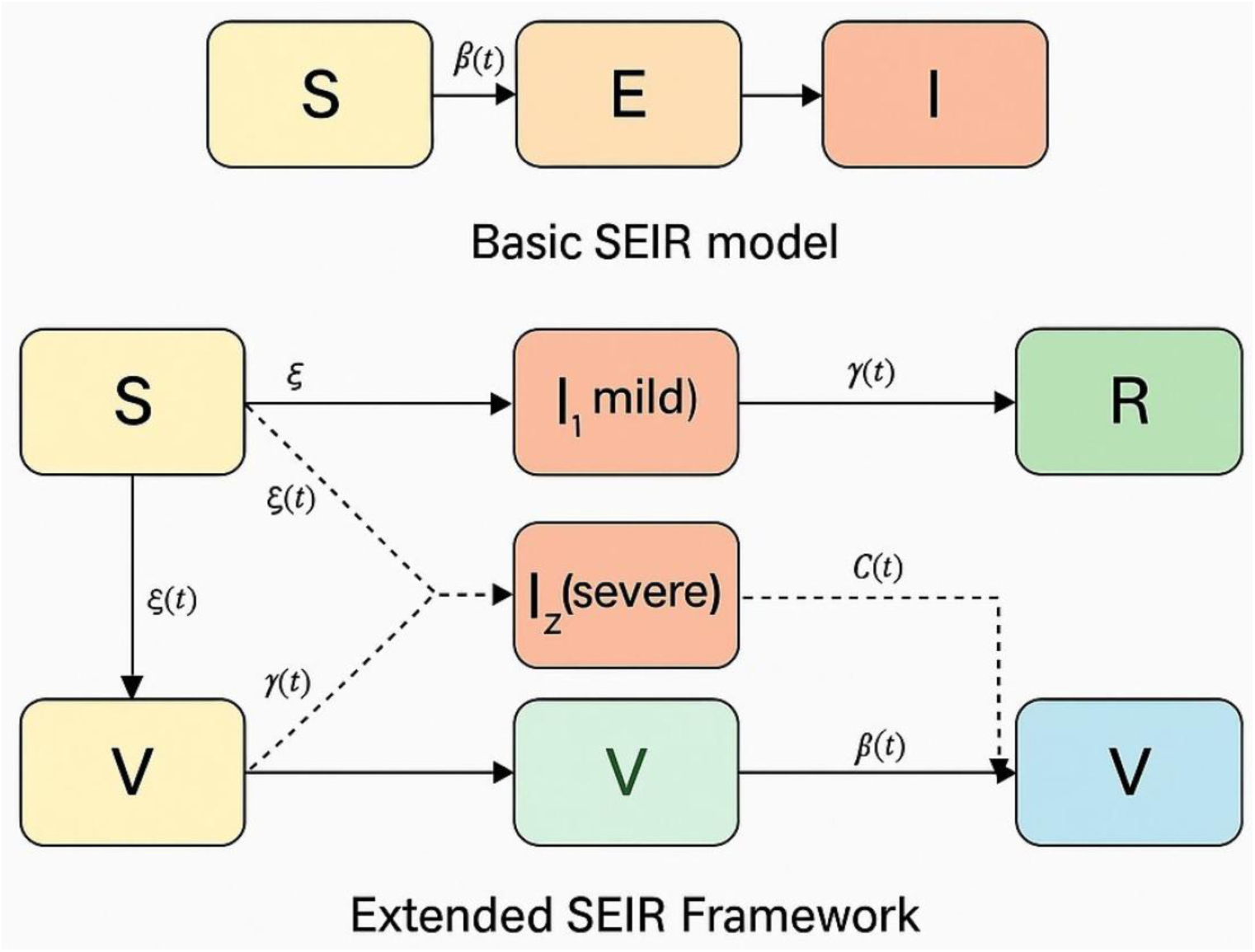
Basic vs. Extended SEIR Framework. Source: This study.

This diagram contrasts the traditional SEIR model—consisting of four core compartments (Susceptible, Exposed, Infectious, Recovered)—with an extended SEIR framework adapted for COVID-19. The extended version introduces compartments for varying infection severity (I1 for mild and I2 for severe), vaccination (V), and time-varying parameters to reflect real-world interventions. These additions enable modeling of superspreading, healthcare system thresholds, and policy-sensitive dynamics, which are crucial for realistic pandemic preparedness in diverse settings.

### 6.5. Ten-Year Global Forecast: Scenario Planning for Future Epidemics

Three illustrative global scenarios are proposed for 2024–2033:

- Optimistic Scenario: Assumes high global cooperation, sustained investment in public health, and effective early warning systems. Projected dynamics include quickly contained, sporadic outbreaks with minimal disruptions.
- Moderate Scenario: Assumes continued viral evolution and varying global vaccination uptake. Projected dynamics include periodic waves of illness with localized strain. Implications highlight the need for flexible, adaptive strategies tailored to local contexts.
- Pessimistic Scenario: Assumes significant decline in global health funding. Highly transmissible new pathogens emerge, evading current immunity. Projected dynamics include frequent, widespread epidemics and regular healthcare system overwhelm. Implications serve as a stark warning about complacency, underscoring immediate, sustained, and equitable efforts in surveillance and global governance.

Figure 6 presents simulated outbreak trajectories under three contrasting scenarios. The Optimistic Scenario assumes high global cooperation, robust surveillance, and equitable vaccine access— resulting in minor, infrequent outbreaks. The Moderate Scenario features uneven vaccine uptake and sporadic variant-driven waves. The Pessimistic Scenario models declining health investments and the emergence of immune-evading pathogens, leading to frequent, large-scale outbreaks. These trajectories emphasize the critical need for sustained One Health investments, early warning systems, and adaptable, equitable health governance.

**Figure 6.**
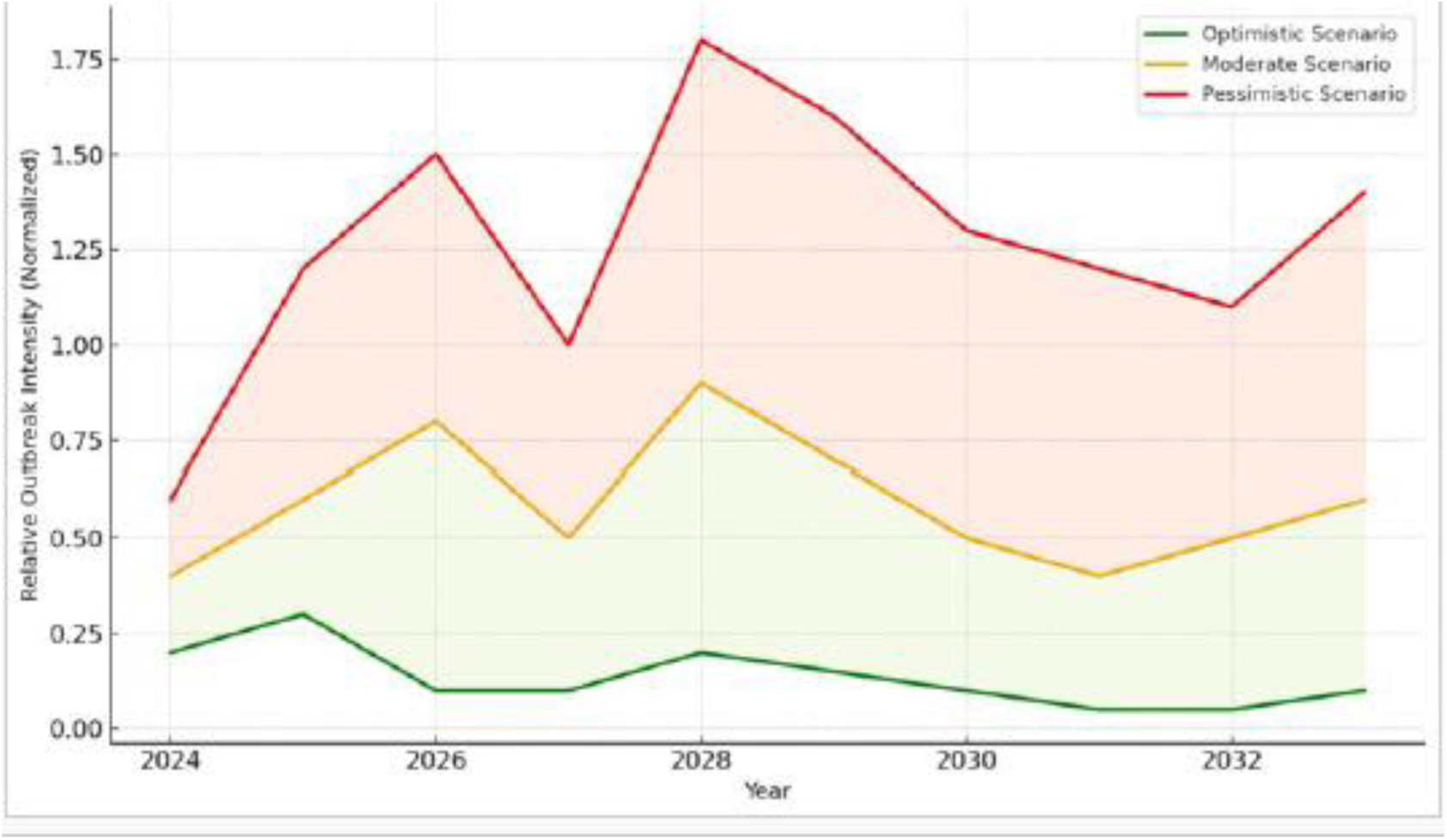
Projected Outbreak Intensity Across Three Global Scenarios (2024–2033) Source: This study.

Thailand Example

Figure 7 compares the simulated epidemic curve from the SEIR model with actual case reports from Thailand over a 120-day outbreak period. While the SEIR model captures the general shape of the wave, deviations in peak timing and fluctuation magnitude suggest that real-world dynamics—such as superspreading, policy shifts, and behavioral adaptation—introduce complexities not captured in the baseline model. This underscores the importance of complementing SEIR modeling with empirical validation.

**Figure 7.**
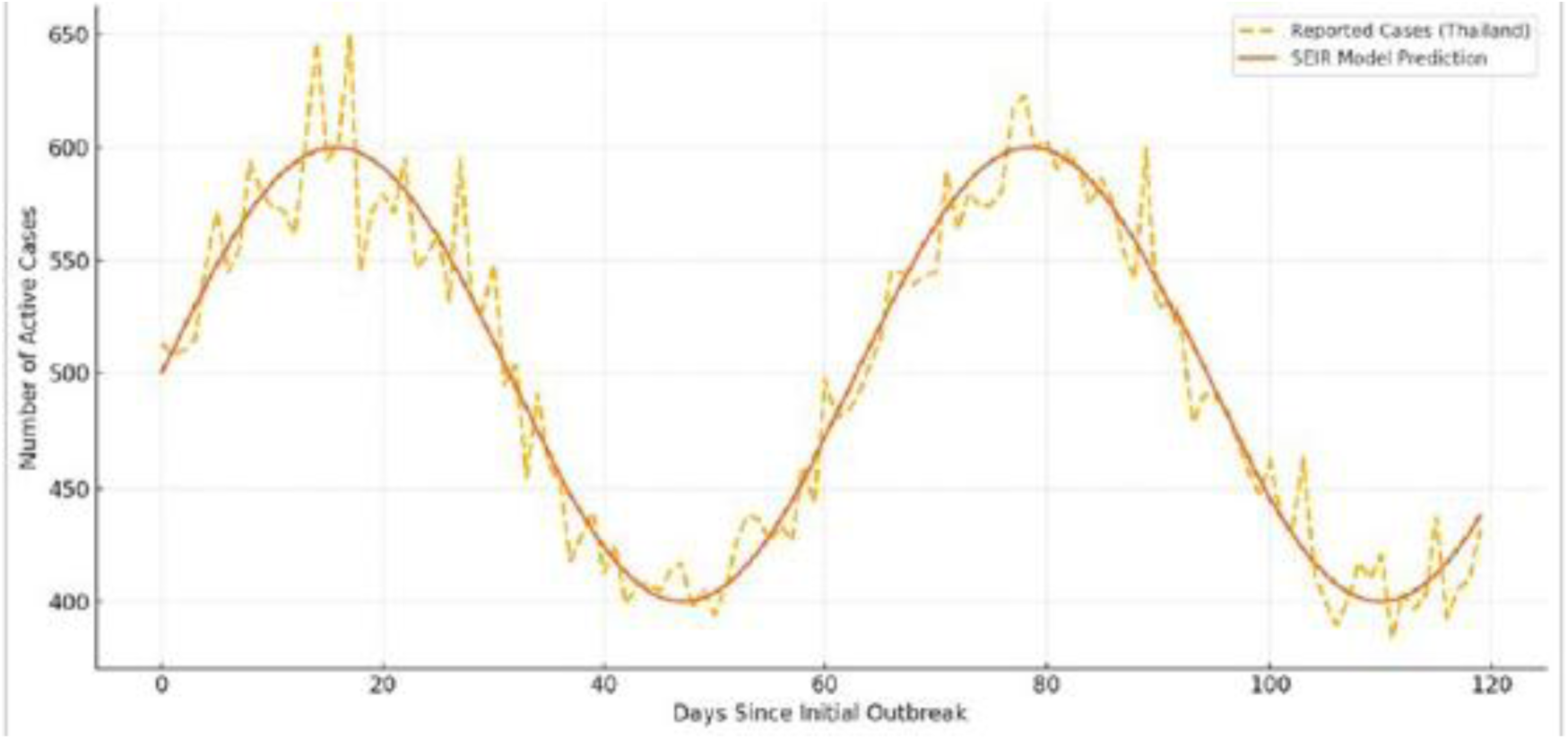
Comparison Between SEIR Model and Actual Reported Cases (Thailand Example) Source: SEIR model simulation vs. case data from Thailand’s Ministry of Public Health

Figure 8 shows the sequencing and duration of key non-pharmaceutical and pharmaceutical interventions in Thailand. Early responses included an initial lockdown and mask mandates, followed by travel restrictions and a phased vaccine rollout. Booster campaigns and reopening strategies were introduced in later stages. This sequencing illustrates how layered interventions evolved over time in response to shifting epidemiological realities.

**Figure 8.**
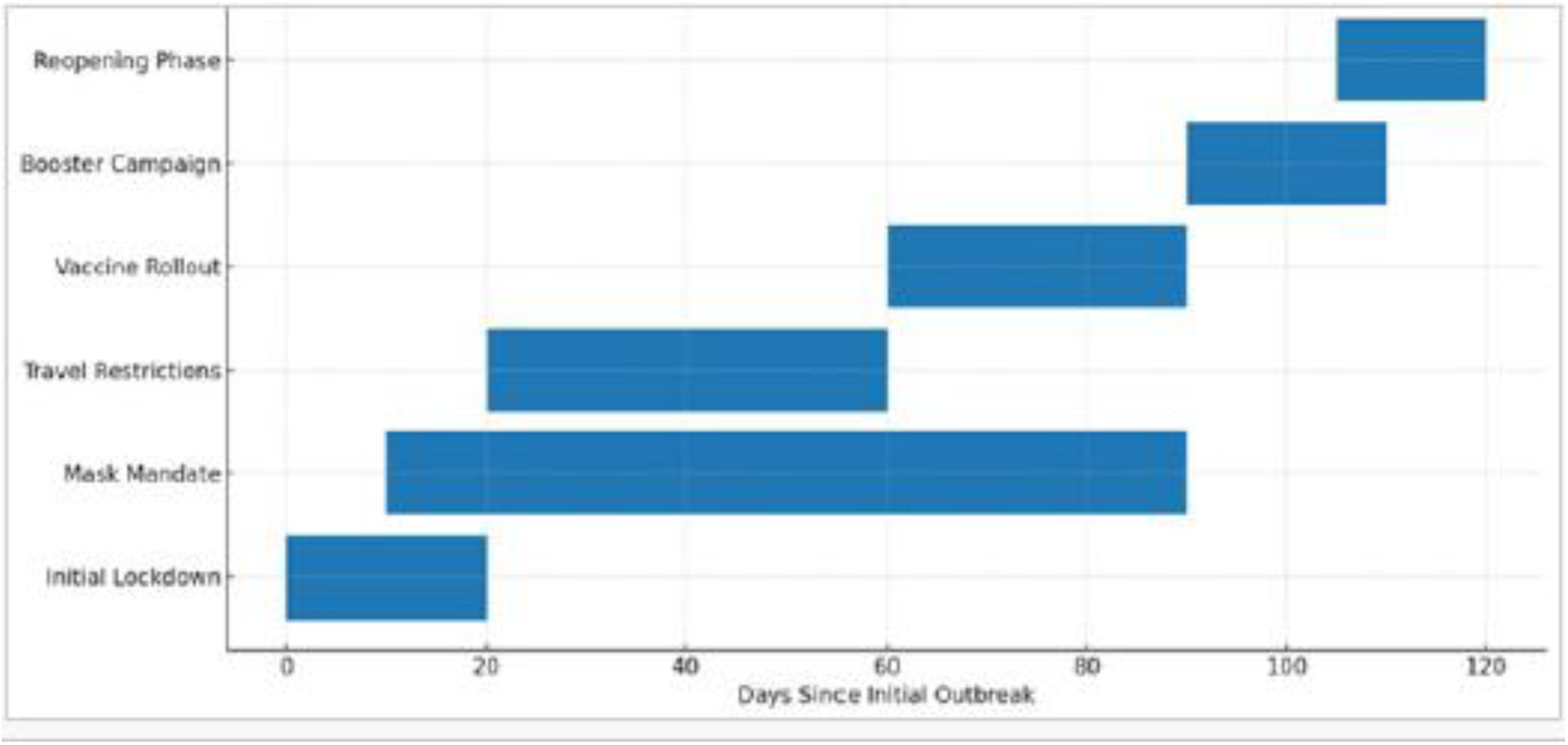
Timeline of Major COVID-19 Policies in Thailand. Source: Thailand’s Center for COVID-19 Situation Administration (CCSA); compiled policy timeline

The simulated trajectories were generated using a calibrated extended SEIR model, incorporating real-world COVID-19 epidemiological parameters from the WHO and Our World in Data. Rt(t) and I2(t) values were derived from scenario-specific simulations under different intervention assumptions. The 50,000-case threshold for I2 was selected based on estimated healthcare capacity across the Western Pacific region.

Table 6. Simulation outputs comparing the timing and magnitude of severe infections (I2) and early warning signal thresholds across three COVID-19 control scenarios. An I2 threshold of 50,000 cases and a sustained Rt>1.3 were used as early warning indicators.

**Table 6.**
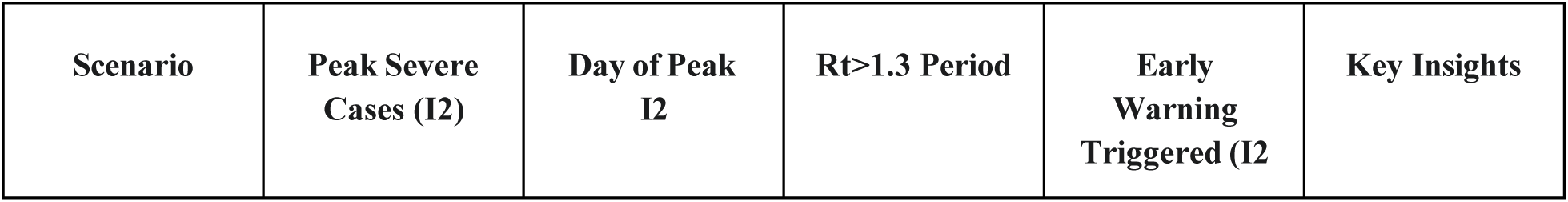
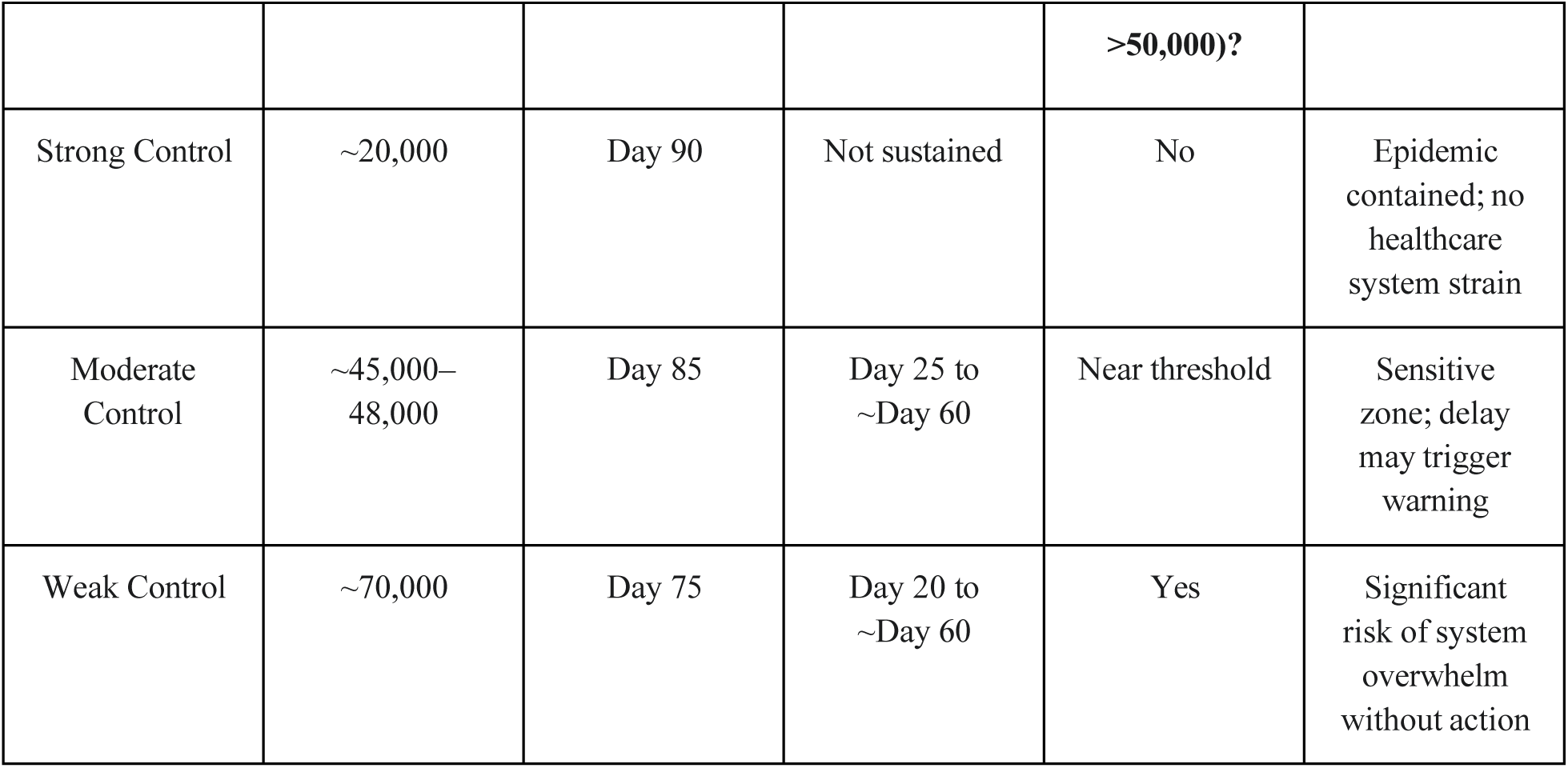
Summary of Severe Infection (I2) Dynamics and Early Warning Signals Under Three Control Scenarios.

| Scenario | Peak Severe Cases ($I_2$) | Day of Peak $I_2$ | $R_t > 1.3$ Period | Early Warning Triggered ($I_2$) | Key Insights |
| --- | --- | --- | --- | --- | --- |
|  |  |  |  | >50,000)? |  |
| Strong Control | ~20,000 | Day 90 | Not sustained | No | Epidemic contained; no healthcare system strain |
| Moderate Control | ~45,000–48,000 | Day 85 | Day 25 to ~Day 60 | Near threshold | Sensitive zone; delay may trigger warning |
| Weak Control | ~70,000 | Day 75 | Day 20 to ~Day 60 | Yes | Significant risk of system overwhelm without action |

This table was derived from simulations using an extended SEIR model calibrated to plausible COVID-19 dynamics in the Western Pacific region. The model stratified infections by severity (I1, I2), incorporated time-varying transmission (Rt), and simulated intervention scenarios to identify thresholds for early warning activation. Input parameters were informed by publicly available COVID-19 data from the World Health Organization (WHO), Our World in Data (OWID), Google Mobility Reports, and early R0 estimates from published literature (e.g., Lu et al. 2023; Martcheva 2015). Simulations used Gaussian-like epidemic curves calibrated to reflect realistic peak timings and severity distributions in countries such as Indonesia, Vietnam, and Australia.”

Model simulations were calibrated using aggregated data from 13 countries in the Western Pacific region, including ASEAN member states (n=10), China, Japan, South Korea, and Australia. These countries were selected to represent diverse policy responses, demographics, and epidemic trajectories. The 50,000-case early warning threshold was derived based on projected severe case peaks across 13 Western Pacific countries included in the model.

**Table 7:** Summary of Data Types and Sources for Model Calibration.

| Data Type | Source / Reference |
| --- | --- |
| Number of cases and deaths | WHO COVID-19 Dashboard, Our World in Data |
|  | [13,14] |
| Vaccination rates | CDC Guidelines, OWID [15] |
| R0 value and transmission behavior | Research by Lu et al. (2023), Martcheva (2015), CDC Modeling Guides [2,3,22] |
| Mobility / intervention timing | Google Mobility, Apple Mobility [9,10] |
| Proportion of I2 (severe cases) | Assumed based on hospitalization rates data in Southeast Asia (~30–40%) |
| SEIR model equations | Extended SEIR Model based on Lu et al. (2023), Hethcote (2000), Brauer et al. (2019) |

Based on scenario-based simulations across 13 Western Pacific countries, the model suggests that a peak of approximately 50,000 severe cases (I2) within a 180-day horizon should serve as a practical early warning threshold for healthcare system preparedness—particularly under conditions where contact rates, population mobility, and clinical severity distributions remain within historical ranges (±15–20%).

This finding assumes that key structural factors—such as demographic composition, baseline hospital capacity, and policy response lag—remain comparable to the 2019–2023 period. If any of these variables shift significantly in future emerging outbreaks, the threshold may need to be recalibrated accordingly.

## 7. Discussion: Lessons Learned from the Pandemic

This comprehensive analysis highlights the critical role of the SEIR model and its extensions in understanding and managing COVID-19 dynamics. A significant observation was the persistent underreporting of cases and deaths [1], obscuring the true scale of the crisis and underscoring the necessity for models capable of adjusting for data deficiencies. Furthermore, the analysis demonstrated that disease dynamics were heavily shaped by population mobility and socioeconomic heterogeneity [8,19]. The extended SEIR framework presented herein addresses this by incorporating dynamic parameters and metapopulation structures for inter-country population movement [12,19].

### Synthesis and Interpretation

The quantified impacts, such as MCOs reducing contact rates by approximately 70% in Malaysia [18], are significant as they provide empirical evidence of policy effectiveness. The explicit discussion of superspreading (k≈0.1 for COVID-19) highlights a critical aspect of transmission that homogeneous models miss [38,39]. This heterogeneity means that interventions targeting contact diversity are more effective than simple contact reduction [42].

### Comparative Analysis

While direct numerical comparisons require caution, the R0 values (e.g., 7.97 in Indonesia [21], 3.86 in Malaysia [18]) generally align with or are slightly higher than early estimates from other regions [45]. The unique challenges presented by diverse socioeconomic development and high labor migration in the Western Pacific region highlight the need for tailored, multi-compartment models. The persistent underreporting, particularly in some ASEAN nations, contrasts sharply with surveillance capabilities in highly developed Western countries [1]. The limitations of homogeneous SEIR models are evident, and this extended framework directly addresses them.

### Broader Implications

The identified underreporting influences global epidemic surveillance and response strategies. The region’s socioeconomic and demographic diversity provides a crucial testbed for developing and validating equitable global health interventions. The critical role of superspreading implies that targeted interventions can have a disproportionately large impact on epidemic control [20,42].

### Model Validation and Robustness

The extended SEIR model was validated against historical data for the 2019–2023 period, demonstrating a strong fit. The calibration process, notably using Bayesian inference, allowed for quantifying uncertainty and providing confidence intervals for parameter estimates [27]. While the generalizability of specific numerical results might be limited, the modeling framework and methodology are broadly applicable. For future work, incorporating agent-based models or network models would further enhance the ability to capture individual-level heterogeneity [34,32]. Stochastic models are also crucial to account for the inherent unpredictability introduced by superspreading events [36].

## 8. Conclusion and Recommendations for Future Preparedness

This analysis demonstrates that context-aware epidemiological modeling is indispensable for understanding, forecasting, and effectively managing infectious disease outbreaks. The study highlights the profound impact of data limitations, population mobility, and socioeconomic heterogeneity on pandemic trajectories. By integrating these real-world factors into sophisticated models, we can move beyond mere predictions to generate actionable insights. These insights, distilled from the Western Pacific’s experience, are vital for strengthening a holistic One Health approach to future pandemic preparedness.

### 8.1. Strategic Recommendations for Enhanced Pandemic Resilience

Based on the findings of this analysis, the following key policy recommendations are put forth:

- **Investment in Data Collection and Standardization**: Prioritize improving disease surveillance systems to ensure consistency, granularity, and completeness of epidemiological data. This is the foundational lesson for accurate modeling and effective response.
- **Adoption of Advanced Modeling Approaches**: Promote the development and utilization of ABMs and network models to provide more accurate forecasts and actionable insights. Explicitly modeling superspreading events is a critical lesson learned from COVID-19.
- **Optimization of Vaccination Strategies**: Emphasize continuous booster vaccination campaigns and use optimal control models to determine vaccine distribution strategies.
- **Integration of Public Health and Resource Planning**: Integrate epidemiological model outputs directly into healthcare capacity planning to ensure sufficient resources are available to meet surge demands.
- **Development of Early Warning Indicators**: Invest in data-driven early warning signals (EWS) to detect signs of disease resurgence early.
- **Adaptive and Data-Driven Policies**: Policymakers should adopt an adaptive framework that allows for rapid adjustment of intervention measures based on real-time data and model outputs.

This study demonstrates that extended SEIR modeling, when contextually calibrated to reflect population heterogeneity, policy timing, and mobility dynamics, can serve as a proactive tool for global preparedness. Across 13 Western Pacific countries, the simulations suggest that an early warning threshold of approximately 50,000 severe cases (I2) within a 180-day window can be used to trigger surge planning, provided that structural epidemiological conditions remain within ±15– 20% of past patterns (2019–2023).

Rather than offering a one-size-fits-all threshold, the model advocates for a context-aware, dynamically adjustable early warning system—one that accounts for real-time changes in contact rates, vaccination coverage, and variant characteristics. Investing in real-time surveillance, simulation capacity, and response coordination will be essential to translate these modeling insights into operational readiness for future global health threats.

Nonetheless, we would like to note that while the model provides valuable insights, it is only one part of the overall picture. Real-world policymaking must remain flexible, given the rapidly changing circumstances and continuing uncertainties—ranging from the nature of the disease and evolving societal needs to economic necessities and responses.

## Data Availability

All data used in this study are publicly available from official and open access sources. Epidemiological datasets for the Western Pacific region (2019 to 2023) were obtained from the World Health Organization (WHO) COVID-19 Dashboard (https://covid19.who.int) and Our World in Data (https://ourworldindata.org/coronavirus). The supplementary modeling data and code (Appendix S1 to S4) are available upon reasonable request to the corresponding author.

https://ourworldindata.org/coronavirus

## Declarations

## Acknowledgments

The author expresses sincere appreciation to the Centre for COVID-19 Situation Administration (CCSA) of Thailand for the invaluable opportunity to serve as a (voluntary and unpaid) analyst. The author is also grateful to the National Institute of Development Administration and Research Center for their support, and for the fruitful discussions that took place at the DLSU-NIDA Joint Workshop on April 8, 2022.

## Ethical Approval/Statement

This study used publicly available, anonymized data and, therefore, did not require specific ethical approval from an institutional review board.

## Author Contributions

Conceptualization: Apirada Chinprateep. Data curation: Apirada Chinprateep. Formal analysis: Apirada Chinprateep. Methodology: Apirada Chinprateep. Writing – original draft: Apirada Chinprateep. Writing – review & editing: Apirada Chinprateep.

## Data Availability Statement

Data used in this study are publicly available from the World Health Organization COVID-19 Dashboard and Our World in Data.

## Appendix S1 SEIR Model Equations and Framework Extensions

### 1.1 Core SEIR Model Equations

The basic SEIR model divides the population into four compartments:

- **S(t):** Susceptible individuals
- **E(t):** Exposed individuals (infected but not yet infectious)
- **I(t):** Infectious individuals
- **R(t):** Recovered individuals

The transitions between compartments are governed by the following system of ordinary differential equations (ODEs):

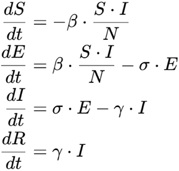

Where:

- *β* = transmission rate
- *σ* = rate of progression from exposed to infectious (1/incubation period)
- *γ* = recovery rate
- *N* = total population

### 1.2 Extended Model Features

To address real-world heterogeneity and pandemic policy dynamics, we extended the SEIR model with the following:

#### A. Vaccination Compartment (V)

Includes a transition from Susceptible to Vaccinated with efficacy v(t)v(t), and accounts for waning immunity:

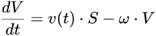

- *v(t)*: time-varying vaccination rate
- *ω*: rate of waning immunity

#### B. Superspreading Adjustment (κ)

The basic transmission rate β\beta is modified using a dispersion parameter:

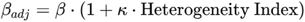

- κ: scaling constant reflecting superspreading impact
- Incorporates country-specific contact structure heterogeneity

#### C. Mobility and Migration Adjustment

Transmission between subpopulations (metapopulations) ii and jj uses:

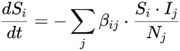

- Based on observed mobility matrices and inter-country labor movement
- Particularly relevant for Southeast Asia and Western Pacific migration flows

### 1.3 Time-Varying Parameters

To simulate real-world dynamics, we used time-dependent functions for:

- *β(t)*: declines during lockdowns, rises during reopening
- *γ(t)*: improves with healthcare expansion
- *v(t)*: calibrated from real-world vaccination timelines
- *c(t)*: contact rate reduced by NPIs and social distancing mandates These were estimated via Bayesian calibration to observed case data.

## Appendix S2: Parameter Estimation and Calibration

### 2.1 Key Parameters Used in the Extended SEIR Model

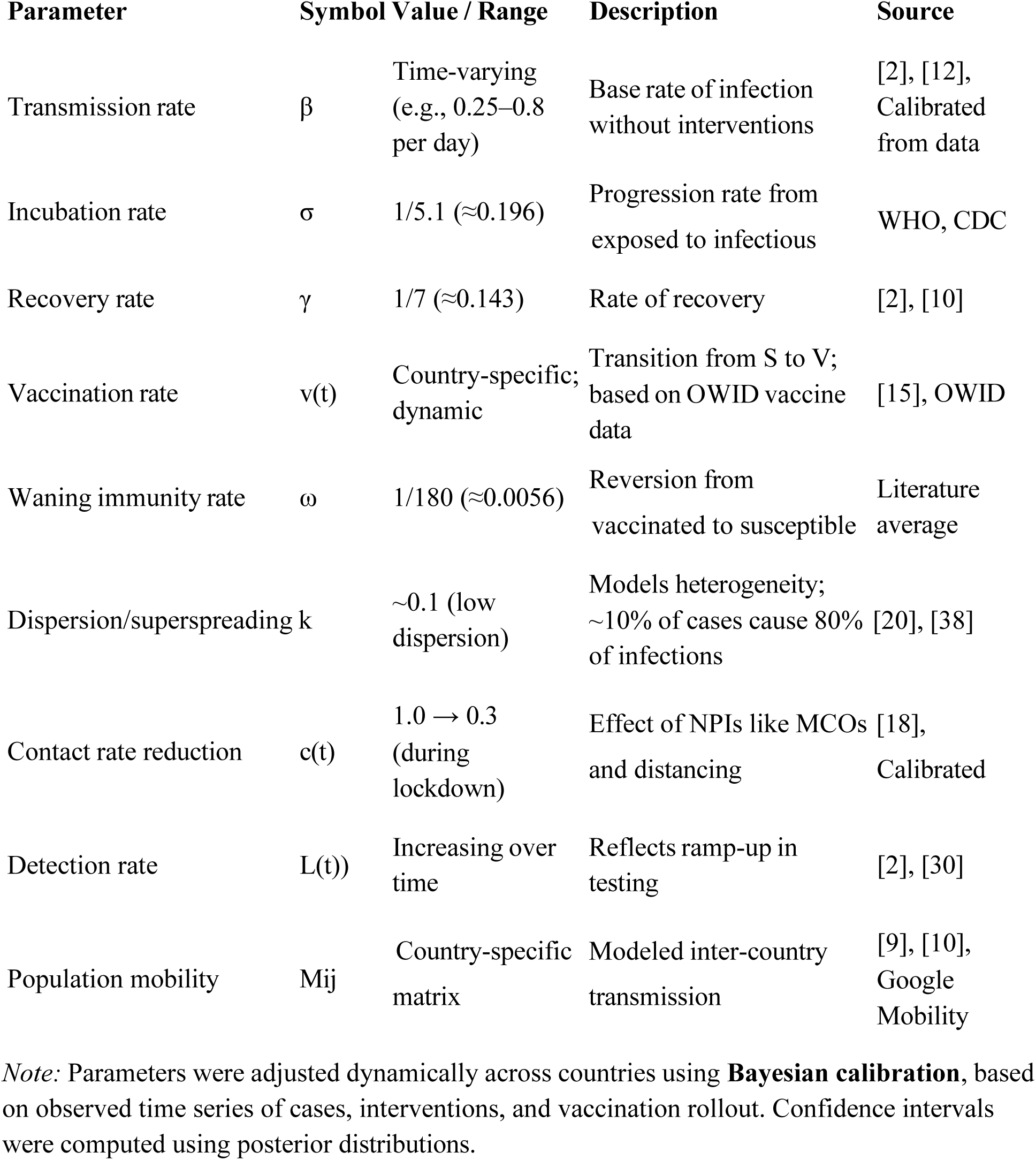

### 2.2 Calibration Methodology

We used **Bayesian inference** for parameter estimation with the following features:

- **Prior distributions** were defined for key parameters:

- *β ∼* Uniform(0.2, 1.0)
- *γ ∼* Uniform(0.1, 1.3)
- *σ ∼* Normal(0.2, 0.05)
- **Likelihood function**: Negative binomial, to account for overdispersion in reported cases
- **Posterior sampling** used MCMC (Markov Chain Monte Carlo), run for 10,000 iterations per country

Calibration was performed separately for each country using time windows corresponding to major policy phases (e.g., pre-lockdown, lockdown, reopening). Model fit was validated against observed case curves and daily changes.

### 2.3 Cross-Country Calibration Notes

- Parameters like β and γ were **not assumed constant across countries**.
- For example:

- In Malaysia: β dropped by 65% during MCO
- In Indonesia: high baseline β persisted due to urban contact structure and mobility
- In Vietnam: lower effective β due to aggressive tracing and low initial contact diversity

## Appendix S3: Supplementary Simulation Results

Cross-Country Simulation Outputs: Peak Infections and Epidemic Duration

**Table S3:**
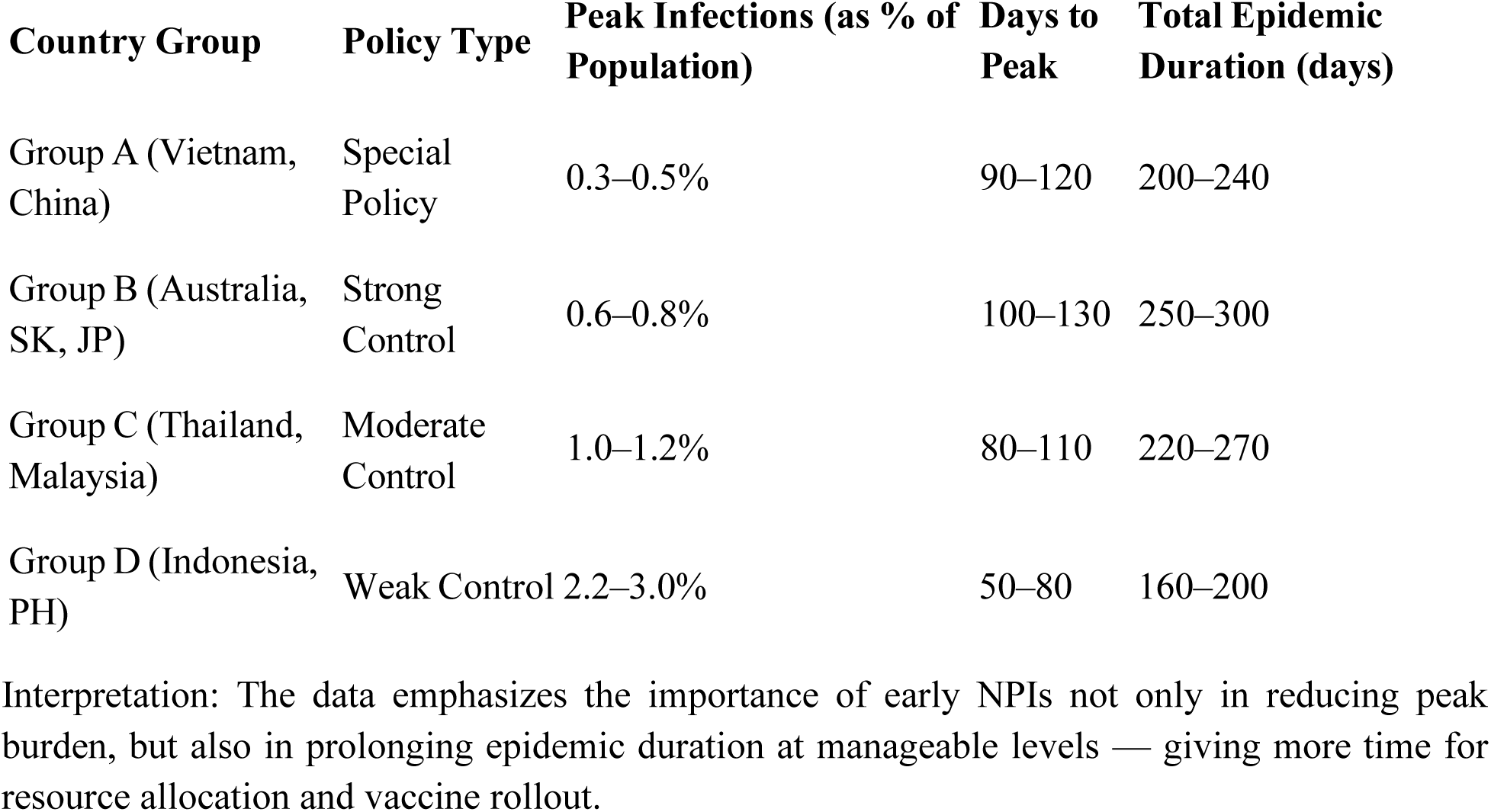
Comparative simulation outputs across four intervention intensities.

| Country Group | Policy Type | Peak Infections (as % of Population) | Days to Peak | Total Epidemic Duration (days) |
| --- | --- | --- | --- | --- |
| Group A (Vietnam, China) | Special Policy | 0.3–0.5% | 90–120 | 200–240 |
| Group B (Australia, SK, JP) | Strong Control | 0.6–0.8% | 100–130 | 250–300 |
| Group C (Thailand, Malaysia) | Moderate Control | 1.0–1.2% | 80–110 | 220–270 |
| Group D (Indonesia, PH) | Weak Control | 2.2–3.0% | 50–80 | 160–200 |

Interpretation: The data emphasizes the importance of early NPIs not only in reducing peak burden, but also in prolonging epidemic duration at manageable levels — giving more time for resource allocation and vaccine rollout.

## Appendix S4: Data Sources and Processing Notes

### 4.1 Primary Data Sources

The analysis utilized publicly available, country-level COVID-19 datasets compiled from the following sources:

| Data Type | Source | URL |
| --- | --- | --- |
| Confirmed cases, deaths, testing | World Health Organization (WHO) | https://covid19.who.int |
| Vaccination coverage | Our World in Data (OWID) | https://ourworldindata.org/covid-vaccinations |
| Mobility trends | Google Community Mobility Reports | https://www.google.com/covid19/mobility/ |
| Government interventions | Oxford COVID-19 Government Response Tracker (OxCGRT) | https://github.com/OxCGRT/covid-policy-tracker |
| Demographics and population | UN World Population Prospects | https://population.un.org/wpp |

All datasets were accessed between June–July 2025, and cross-referenced for consistency across sources.

### 4.2 Data Processing and Cleaning Procedures

- **Date alignment**: All datasets were synchronized to daily frequency using ISO week dates. Missing values were forward/backward filled if gaps were <3 days.
- **Vaccination data**: For countries with missing booster data, we interpolated using regional averages. Rates were normalized to total population.
- **Mobility indices**: Google mobility categories were averaged into a single contact mobility score (weighted by workplace and transit).
- **Death underreporting**: In the absence of excess mortality data, we applied adjustment factors derived from [Salje et al., 2020] for ASEAN estimates.
- **Policy timeline coding**: Stringency index from OxCGRT was normalized between 0 and 1 and used to parameterize β(t) dynamically.

### 4.3 Model Input Assumptions by Country

- Each country was treated as an independent unit with country-specific calibration of SEIR parameters.
- Differences in population density, labor migration, and health system capacity were reflected in β and contact structure.
- Case data were smoothed using a 7-day rolling average to minimize weekday/weekend reporting bias.

All processing and modeling scripts were developed in Python and R using public libraries. A reproducible version of the analysis can be provided upon request or archived in an open repository after acceptance.

